# Improving the Induction of PPI Contributors on Trial Oversight Committees

**DOI:** 10.1101/2020.04.20.20054106

**Authors:** Emily C Pickering, Bec Hanley, Philip Bell, Jacqui Gath, Patrick Hanlon, Robert Oldroyd, Richard Stephens, Conor D Tweed

## Abstract

Clinical Trials Units are encouraged to integrate Patient and Public Involvement (PPI) into all aspects of trial design, delivery, oversight and dissemination. This research explored the induction and training of PPI Contributors joining trial oversight committees. It was used to create an induction pack for new PPI Contributors at the Medical Research Council Clinical Trials Unit at University College London’s (MRC CTU at UCL). We have made this resource available to all researchers and in this we report describe the methodology behind its production.

---

In the United Kingdom, standard practice for clinical trial oversight and management is tripartite in line with European Medicines Agency guidelines^1^, being formed of a Trial Steering Committee (TSC), Trial Management Group (TMG) and an Independent Data Monitoring Committee (IDMC)^2^. In recent years, greater importance has been placed on PPI within clinical trials^3-6^. Many funding bodies now insist upon completed and planned PPI activity being detailed in grant applications^7-10^, and under the incoming European Union Clinical Trials Regulation any public engagement will have to be documented in study protocols^11^. In addition, the 2018 National Standards for Public Involvement from the National Institute for Health Research^12^ emphasise that PPI best practice involves partnership throughout the life of a study’s design, running and governance. The inclusion of one or more PPI Contributors on trial oversight groups is one way to address the need for public involvement in trials at the highest level of oversight and accountability. However, sitting on a trial oversight committee requires patient/public members to have skills and understanding beyond the experiential contributions that are more common in PPI generally.

Training materials for PPI Contributors in clinical research have been found to be of poor quality^13^ despite evidence showing that involvement is most valuable when Contributors are adequately supported and have clear roles and responsibilities^14^. Clear guidance and better support from the outset of their involvement in a clinical trial is therefore required. The Medical Research Council Clinical Trials Unit at UCL (MRC CTU at UCL) aims to have PPI Contributors on all trial oversight committees as per their committee charters^15^. However, the last induction pack for the unit was developed in 2011 and it was agreed to create a wholly new pack. In collaboration with PPI Contributors experienced in trial oversight work, we identified the requisite topics that should be covered in an Induction Pack and created an updated resource for the MRC CTU at UCL, encompassing current best practice and research findings.

## Methods

A literature search of published articles detailing the requirements and methods for training and induction of new PPI Contributors was performed. Requests for sharing unpublished induction materials were sent to CTUs and charities with an interest in PPI in research. Key themes and topics found from these materials were then triangulated through identification of repeated concepts and ideas from a variety of sources. These then formed the initial sections of a draft Induction Pack presented to a workshop of existing PPI Contributors from the MRC CTU at UCL. A revised draft pack was based on the feedback from the PPI workshop alongside incorporating andragogical theory and best practice. A second review by workshop attendees resulted in the completion of a final template.

### Sample

Based on the known breadth of synonymous terms for PPI and trial oversight committees, a wide-ranging search strategy of Embase, MEDLINE and PsychINFO was employed to identify suitable literature for review. The search strategy employed is listed in Table 1. Inclusion criteria were references to:

**Table 1:**
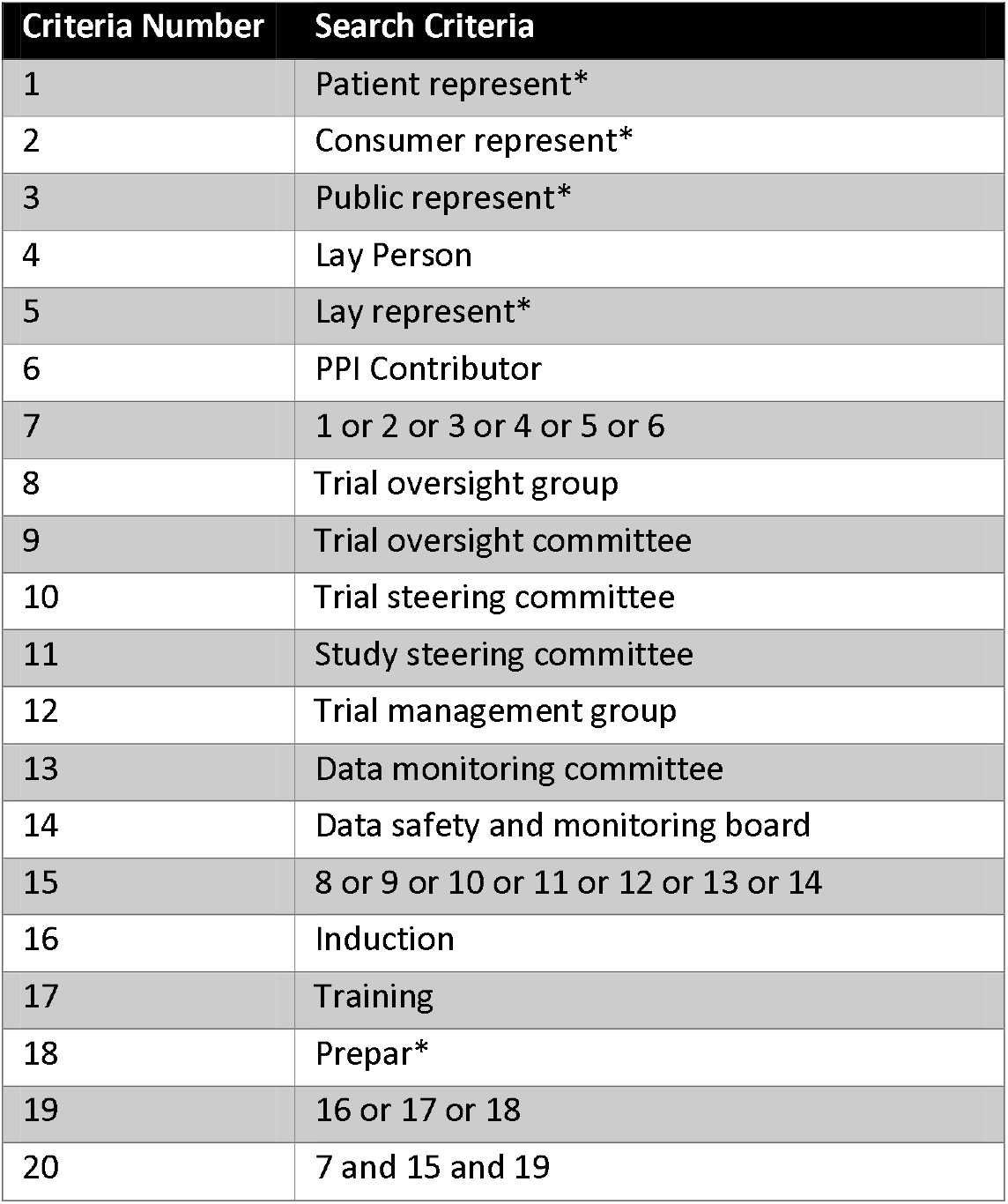
Published data search strategy

1. training or induction materials
2. at least one type trial oversight committee (TMG, TSC or IDMC)
3. PPI Contributors

PPI leads at CTUs were approached to share local induction materials via the UK Clinical Research Collaboration’s Registered Clinical Trial Units Network (UKCRC CTU). This network of CTUs was selected as these organisations are recognised as experienced centres in clinical trial delivery and design^16^. The network hosts a dedicated PPI sub-group with the specific aim of mapping existing resources and developing collaborative working^17^.

Two requests, in April and May 2018, to share information were sent to research-focused charities via the Charities Research Involvement Group – an organisation dedicated to supporting PPI in health research^18^. It was hypothesised that the collaborative aim of this group would result in a greater number of resources being shared than if individual organisations were to be indiscriminately contacted directly. Approaching a collective also allowed for a wide range of organisations to be contacted in an efficient manner.

### Data Collection

#### Published data

Abstracts of articles returned through the database searches were reviewed to assess suitability for inclusion in the results by EP. Those deemed to meet at least two of the inclusion criteria stated above were reviewed in full. Reference lists of the most relevant articles were checked to identify any additional relevant materials using the same approach of abstract review.

#### Unpublished data

CTUs that responded to the request for information sent by the UKCRC and charities involved in the Charities Research Involvement Group were invited to discuss their materials and methods by phone or email or to provide electronic copies of their resources directly. Any materials referenced in the documents that were not captured in the literature search were reviewed in light of the inclusion criteria above.

#### Workshop

Fourteen PPI Contributors were invited to a morning workshop held in July 2018 at the MRC CTU at UCL. A draft Induction Pack, created from the most common themes that had emerged from the literature reviews, was circulated to those attending in advance. Themes were selected based on the frequency in which they were covered in the induction materials gathered from the literature search, with multiple mentions in the same material each counted. The resulting sections of the initial draft Induction Pack were:

- Clinical Trials
  - What is a clinical trial?
  - Why do trials need to be conducted?
- Medical Research Council Clinical Trials Unit at University College London
  - What is the MRC CTU at UCL?
- Trial X
  - What is *Trial X*?
  - What are the objectives of *Trial X*?
  - What will happen during *Trial X*?
  - How are patient representatives involved in *Trial X*?
- Trial Management Group
  - What is the TMG?
  - What is your role on the TMG?
  - How will you support the TMG?
- Trial Steering Committee
  - What is the TSC?
  - What is your role on the TSC?
  - How will you support the TSC?
- Independent Data Monitoring Committee
  - What is the IDMC?
  - What is your role on the IDMC?
  - How will you support the IDMC?

The attendees were divided into two separate groups. The first half of the discussion focussed on the generic sections of the Induction Pack which would be relevant to a member of any oversight committee. The second half of the session then concentrated on details required for new members of either a TMG or TSC. One type of committee was considered by each group, based on the experience of each individual PPI Contributor so as to benefit from direct expertise and first-hand knowledge. Only one attendee had experience of IDMC membership therefore feedback was collected by email following the workshop. Feedback and discussion were collated through individual electronic review after the workshop so that all attendees could participate in the group discussions.

Discussions were facilitated by one PPI Contributor from the MRC CTU at UCL PPI Group and one member of CTU staff, supported by a student note-taker. Attendees were informed that the draft Induction Pack was open to as much change as they deemed necessary. However, the discussions and suggestions had to focus on PPI Contributor needs only. Wider training requirements, such as Induction Packs for researchers or other members of the committees, did not fall within the remit of this research and therefore should not be considered.

#### Individual Review

PPI Contributors were offered the opportunity to conduct a second, independent review of the final draft of the Induction Pack which incorporated the feedback from the workshop and academic theory on adult learning. The document was shared via email and comments were recorded and submitted via tracked changes.

### Data Analysis

An estimate-talk-estimate consensus method was implemented^19^, with the addition of a further feedback-estimate round to complete the Induction Pack review process (figure 1). Key themes and topics covered in the materials collected were extracted and triangulated through selective coding^20^ to identify the most and least common subjects discussed on the topic of training and induction. These data informed the initial sections included in the draft Induction Pack produced for review at the PPI Workshop.

**Figure 1.**
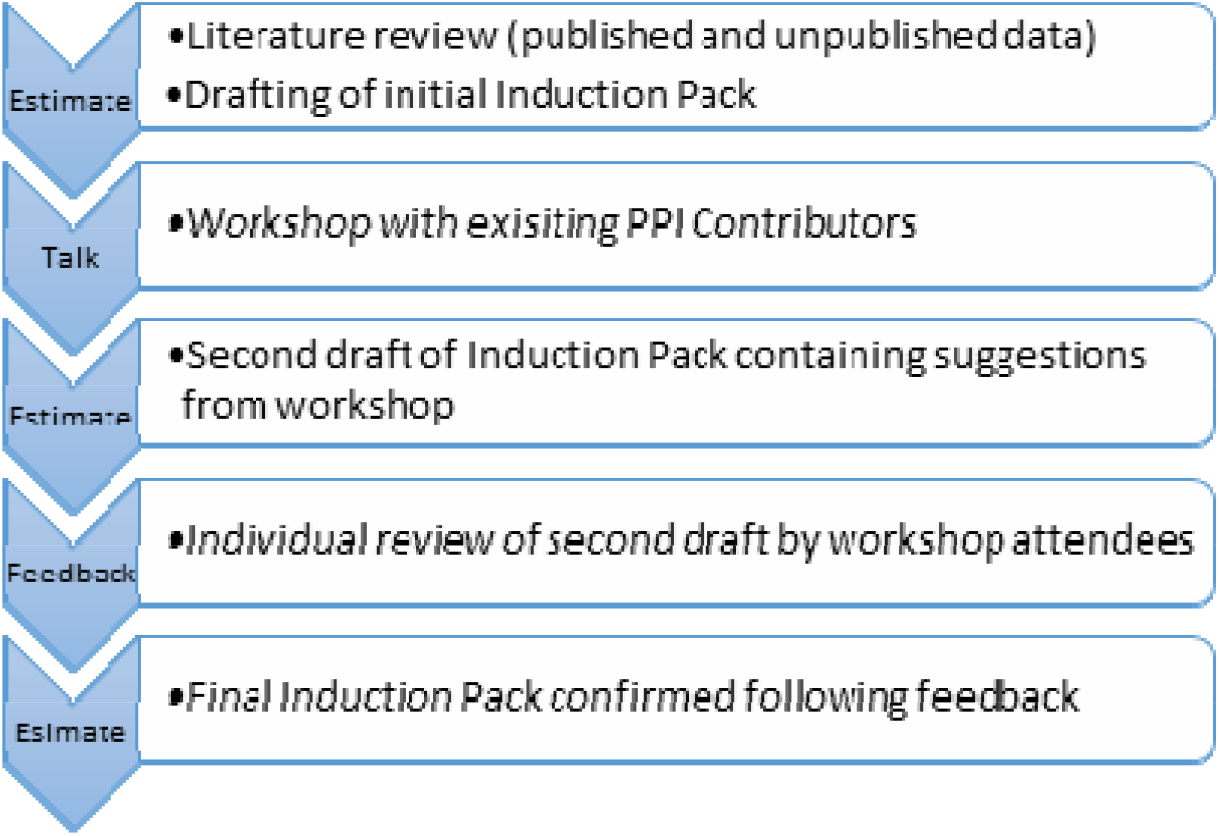
Process of data collection for Induction Pack

Notes taken from the workshop were coded and added to the existing data to confirm suitability of the current sections, identify missing topics felt to be of high importance, suggest language amendments and remove information not felt necessary for inclusion in the Induction Pack.

A final draft of the Induction Pack was subsequently produced, taking into consideration the vetted workshop feedback, and distributed to the workshop participants for a second review. This was completed individually by workshop participants and collated for review before being included in the final pack.

## Results

### Published Data

No published data was found when using the search criteria laid out in the Methods. Several iterations of broader search terms were required before a manageable number of suitable abstracts were returned, but very limited articles merited full review.

Results focused predominantly on the importance of adequate support for PPI Contributors in research. However, no detailed guidance or template was found in any of the results returned. Baxter et al.^21^ conducted a systematic review of PPI training and support in 2016 which did identify some model resources. The studies reviewed focused on involvement in interview panels for new clinicians, wider health research or service reviews. None of the identified studies involved PPI Contributors on clinical trial oversight committees.

The EUPATI (European Patients’ Academy on Therapeutic Intervention) overview^22^ gave a description of the programme and highlighted their toolbox of existing training materials but did not provide any critique of the training^23,24^. Template training programmes are available for researchers under the title “mini-course starter kits” which group together existing resources from within the toolkit to cover certain topics, including DMCs and TSCs ^a^. These are, however, starting points for researchers to adapt and make relevant for their specific trial and parts of the course (e.g. quizzes for each section) are yet to be developed.

Of note, Bagley et al.^25^ highlighted the lack of an induction pack for new PPI Contributors on trial oversight committees in Phase 2 of the creation of their toolkit for PPI in trials. Their finding reinforced the necessity of this piece of research; the existing resources identified by Bagley were reviewed and added as additional data for analysis.

### Unpublished Data

Existing induction materials from five CTUs and three research charities were provided or discussed in addition to the existing MRC CTU at UCL Induction Pack. The topics covered in these materials are summarised in Table 2. In addition, two other CTUs responded to say they did not currently have any localised induction materials but planned to create such resources in the near future. No organisation refused to share their documents.

**Table 2:** Unpublished training material subjects

| Theme/Topic | Frequency |
| --- | --- |
| Role of a PPI Contributor | 11 |
| Explanation of what an oversight committee is | 8 |
| Number of meetings to attend | 8 |
| Who to contact for more information/support | 7 |
| Background to the specific trial under review | 6 |
| Key qualities of a PPI Contributor | 6 |
| Description of the role of each committee | 5 |
| Activity required between meetings | 5 |
| Confidentiality | 5 |
| Payment/expenses | 5 |
| Explanation of what PPI is | 4 |
| Non-attendance of meetings | 4 |
| Who makes up the membership of the committee | 4 |
| Evaluation of involvement (feedback/review) | 4 |
| Examples of activity of Contributors may undertake | 3 |
| Background to clinical trials | 3 |
| What happens at a committee meeting | 3 |
| Duration of the committee membership | 3 |
| Terms of Reference for the committee | 3 |
| Specific training to be completed | 3 |
| Trial timeline (from idea conception to results published) | 2 |
| Implications of involvement on studies (benefits/tax codes etc.) | 2 |
| Location of committee meetings | 2 |
| Training predominantly conducted as a face-to-face discussion | 2 |
| Importance of Plain English | 1 |
| Background to trial design | 1 |
| Common challenges to research | 1 |
| Developing a proposal | 1 |
| Getting your voice heard at meetings | 1 |
| Resignation from the committee | 1 |
| Safety checks to be completed (e.g. DBS) | 1 |
| Application process for committee membership | 1 |
| Explanation of what a CTU is | 1 |
| Glossary of terms | 1 |

### Workshop

The half-day workshop for review of the draft Induction Pack was attended by twelve of the fourteen PPI Contributors invited, all of whom have experience of sitting on trial oversight committees at the MRC CTU. All attendees supported the need for the creation of an improved Induction Pack and discussions from each of the two separate groups provided similar feedback (table 3).

**Table 3:** Feedback from workshop

| To Improve | To Include | To Consider |
| --- | --- | --- |
| **Terminology**<br>•Use "plain English" not "lay language"<br>•Provide definitions of key terms<br>•Make language used more consistent throughout (e.g. standard of care vs current treatment) | **Additional Sections**<br>•Glossary/jargon buster<br>•Signposting for further information (in person or via additional resources)<br>•Supporting/encouraging the distribution of results and implementation into standard practice where possible | **Terminology**<br>•"Patient Representative" is an inappropriate title for some situations (e.g. people with HIV or parents/carers of children)<br>•Clarification of the meaning of "involvement" and "engagement" |
| **Formatting**<br>•Ensure consistency of visual style<br>•Re-ordering of the sections: introduction to trials, then the CTU, then trial specific information | •Advice on getting your voice heard in meetings/teleconferences<br>•Implications of committee membership on personal benefits/taxes (in relation to honorariums) | **Pre-Requisites**<br>•Does the CTU require PPI Contributors to have GCP training?<br>•Indemnity insurance - are PPI Contributors covered by the CTU's policies? |
| **Language choices**<br>•Give clearer description of remit of this role to reduce uncertainty (e.g. remove "not expected to...")<br>•Sentence length is too long for plain English in some sections<br>•Academic language is still used throughout - needs to be simplified | •Encouragement to bring other skills to the group rather than "just" a PPI Contributor<br>•Breakdown of the individual members of the committees and whether they're independent or not<br>•Duration of the committee membership, anticipated workload & how to leave the committee<br>•Access to the protocol and PIS/ICF<br>•Information on what research or trials will have been conducted in the lead up to this study<br>•Why PPI is important<br>•Confidentiality and how to deal with the media | **Layout**<br>•"Pick and Mix" design where trial teams can choose which sections to include<br>•Highlight "essential" and "optional" sections<br>**Future Additional Resources**<br>•Trial-specific guidance on the role and remit of each committee, to be agreed by all members of the committee to ensure consistency throughout the CTU<br>•Future iterations to include multimedia to make the document more accessible (e.g. to those with visual impairments)<br>•Implementation of a "Scientific Mentor" role to provide ongoing |
| **Existing Sections**<br>•Detail the additional visit/testing burden on participants on the study compared to standard of care<br>•Give more details of the aims and objectives of the study under consideration in the trial specific section<br>•Provide links to existing documents and resources rather than creating new material<br>•Include examples of the tasks PPI Contributors may be asked to participate in<br>•Clarify which committees see raw data broken down by treatment group | **Clarification**<br>•Differentiation between the role and remit of each oversight committee<br>•Whether PPI Contributors have voting rights and how they are weighted compared to the other members<br>•Involvement outside of the committee: may be asked to attend conferences to discuss the study<br>•Whether Contributors can join multiple committees | support regarding clinical aspects of the study<br>•Similar induction pack to be created for researchers to assist them in supporting their PPI Contributors<br>•Guidance for pre-trial groups such as protocol review boards<br>**General**<br>•Advise trial teams that a member of the PPI Group reviews the document completed by the Study Team before it's distributed to PPI Contributors<br>•Make it clear to investigators that the acceptable levels of risk taken by patients will vary from group to group<br>•Emphasise that the PPI Contributor's role is to focus on patient safety and wellbeing<br>•Highlight that the role may adapt and change over time as the study progresses<br>•Integrate the NIHR National Standards for PPI<br>**Format**<br>•Bullet points make the text easy to read<br>•It is good to have clear sections marked for trial teams to add in specific data to personalise the pack<br>•Keep the pack as short as possible to avoid overwhelming but provide signposting to additional resources |

The importance of using plain English and short paragraphs for ease of reading was emphasised repeatedly. The differences between the three committees were not always clear to the PPI Contributors, despite many of them having been members of different committees themselves. It was acknowledged that a large amount of information and resources is already in the public domain, often having been created by patient support groups or charities. The workshop attendees felt that it was important to make use of these existing resources rather than “reinventing the wheel”.

### Post-Workshop Review

A second draft Induction Pack was created which assimilated the feedback from the workshop. More attention was paid to style and format, as well as the inclusion of additional key topics and sign-posting to existing materials. Figure 2 shows the amended order of topics. The re-ordering provides a more layered approach to the information presented and allows for new knowledge to be built upon an understanding of earlier concepts thus promoting more critical thinking^26^.

**Figure 2:**
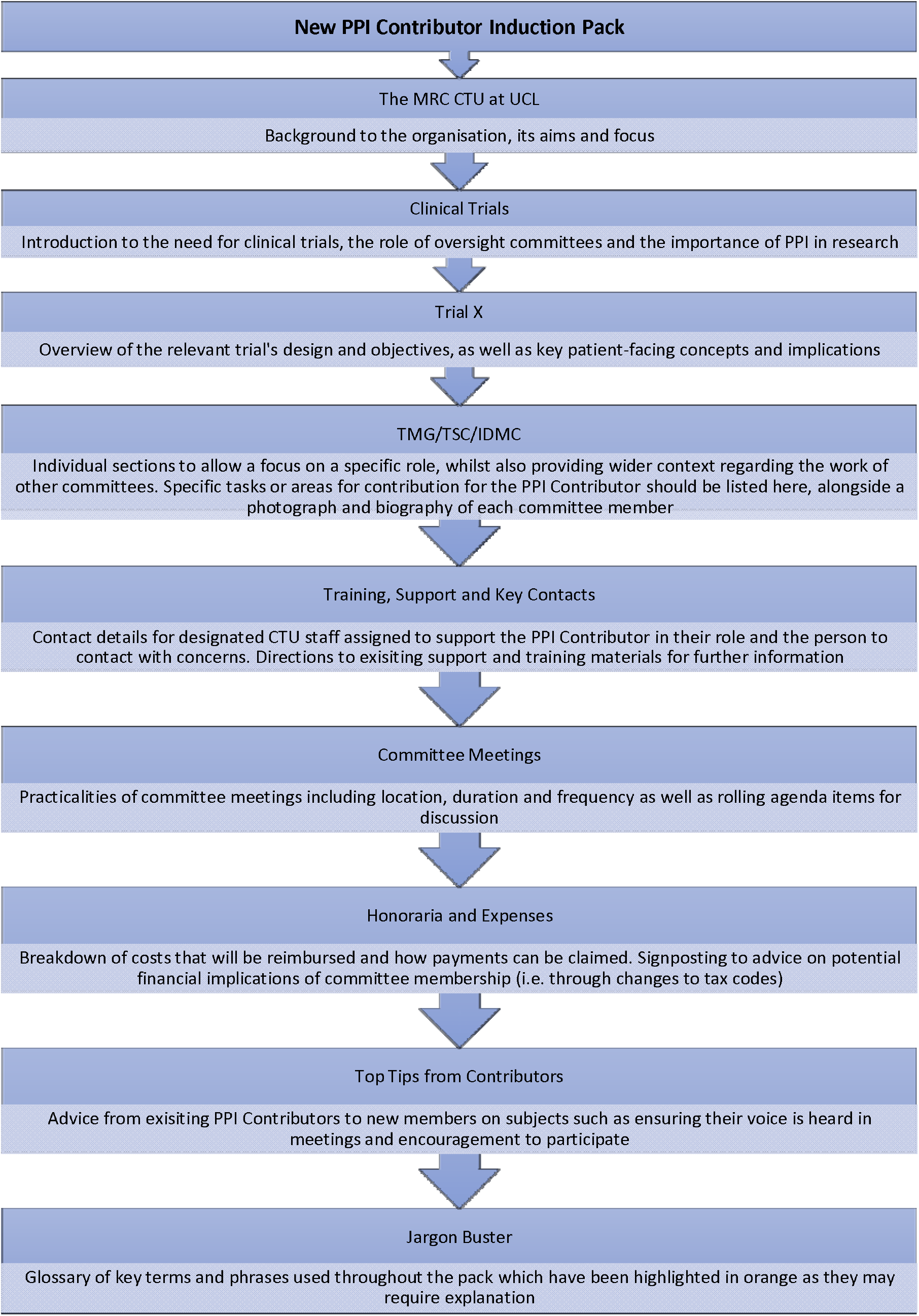
Order of Induction Pack Chapters

Guidance notes for members of trial teams who will be completing the template were added to explain what trial-specific information needs to be provided to personalise the resource. These notes have been written in a different colour and font to distinguish them from the PPI Contributor facing information and trial teams are instructed to delete these notes before the Induction Pack is shared. The Methods and Ethics & Disseminations sections of the SPIRIT Guidelines were additionally used to guide and refine the chapter choices^27^ to make completion of study-specific data as simple as possible. An assumption has been made that trial teams would be very familiar with what information is expected to be provided under these key sections.

### Individual Review

Feedback on the second draft Induction Pack was received from five of the workshop attendees. The majority of the comments focused on the language used and suggested shortening sentences and using more positive and lay-friendly terms. The advice to trial teams was expanded to provide further data or information on the study’s design that was felt to be useful to new PPI Contributors.

More general feedback was also received which did not focus on the Induction Pack itself, but the training and use of PPI Contributors on oversight committees in general. Such comments included:

- Questioning whether the selection of a new PPI Contributor should consider the amount of prior in-depth knowledge of the disease under study held by the candidate to ensure they can adequately represent the patient population
- Highlighting that the majority of the guidance focuses on committees for trials that have already been designed and gained funding. Therefore, it is not suitable for PPI Contributors assisting with regulatory approvals applications and initial protocol writing
- Whether un-equal voting rights between researchers and PPI Contributors should be permitted as this could impair the ability of PPI Contributors to make meaningful changes to the trial
- Encouraging the use of PPI impact assessment sheets which can be completed to track PPI activity and successes on oversight committees
- Details of the law around informed consent
- Creating a Top Tips for Researchers equivalent document
- Advising on the number of PPI Contributors that should be assigned for each committee

## Discussion

This study has found that, despite the uniformity of large aspects of the clinical trial field imposed by regulatory approval processes, international and national standards, and reporting guidelines, there is very little consistency in how patients and the public are involved in trial oversight committees. The lack of published materials on PPI Contributors’ training and involvement in oversight committees has allowed the creation of the Induction Pack template presented here to be very organic and predominantly influenced by direct involvement of experienced PPI Contributors. Workshops with focus groups, such as the one involved in this research, provide primary data from experts on the topic at hand. Discussions are often free flowing and spontaneous, giving participants the opportunity to explore both how and why they feel issues are important^28^. Such qualitative and subjective data is not often found in clinical trials publications. However, its value lies in its insights into the perspectives of the target population^29^.

Since all the attendees at the Induction Pack workshop had first-hand experience of sitting on trial oversight committees, their opinions and experiences provided plenty of data and ideas for analysis which could not be collected from the literature. The pack is therefore representative of the personal experiences of committee members with experience in the field and provides adequate ‘truth value’ to be credible^30,31^. The trustworthiness of the qualitative data collected is further supported by the consistency between the notes taken from the two separate workshop groups.

Whilst different note-takers were used for each of the discussions, neither student disclosed any preconceived assumptions regarding the importance, or otherwise, of any particular topic to be discussed and therefore their records have been considered neutral representations of the conversations held. Further neutrality and completeness would have been achieved had audio recording transcriptions been used for analysis^28^. However, the use of experienced PPI Contributors rather than researchers as the primary facilitators for the focus groups reduced the feeling of distance between the participants and the discussion’s leader, thereby providing a more neutral environment and partnership^30,32^.

### Choice of Chapter Topics

The methodology employed for the collection and analysis of data was carefully considered to ensure the Induction Pack’s applicability and transferability to a wider audience. Coding of both the quantitative literature data and qualitative workshop notes was conducted using the principles of grounded theory^20^: rather than looking to prove or disprove an existing theory using evidence already collected, data was reviewed alongside its collection with the aim of creating new theory. For example, as this research project commenced with the pre-defined goal of creating an Induction Pack, focused selective coding of data related to the topic facilitated the identification of key themes and issues for inclusion in the Induction Pack. It was through this analysis, and the recurrence of key categories, that the final topics for the Induction Pack were chosen therein creating new theory to support the actions taken.

Ideas or comments that were not recurrent, or that could not be grouped into an existing category, or that fell outside the project’s predefined scope were not included in the final resource. The need for complementary guidance for researchers to support new PPI Contributors was raised on several occasions, as was the anticipated benefit of additional ongoing support mechanisms within CTUs for PPI Contributors. Had true grounded theory methodology been implemented – whereby a topic to focus upon is found from the emergent data, rather than seeking evidence related to an idea – these data and concerns might have been further explored. Other research has highlighted the need for improvements in the working relationships between PPI Contributors and academics^33^. However, the objective of this project had been set in advance and there was not scope to explore these ideas further. A potential limitation of this research is highlighted herein, suggesting that wider, or greater, issues of concern to PPI Contributors on oversight committees may have been missed as all analysis had a specific objective in mind. Future iterations could involve a more blue-sky approach with wider objectives.

Two further topics which were relevant to the final objective posed difficulties when deciding whether to include them in the Induction Pack.

The first topic concerned payments for involvement in oversight committees, which has previously been identified as a difficult subject to cover in PPI guidance because it may lead to feelings of inequality of perceived value amongst members^13^. As members of some oversight committees will be attending meetings as part of their salaried role, the difference in reimbursement for time between a CTU staff member and a PPI Contributor receiving an honorarium can be considerable. However, UK guidance from the National Institute for Health Research’s (NIHR) INVOLVE provides clear instruction advocating discussion and agreement of any reimbursement of expenses or payments in advance of PPI activity taking place^34^. The MRC CTU at UCL also has an in-house costing template for all PPI activity to ensure fair recompense for PPI Contributors at all stages of involvement. This topic was, therefore, included in the final Induction Pack.

The second contentious topic was whether to include information on how to withdraw from being a PPI Contributor on a committee. Opinion was split between whether providing this information could discourage new members from fully committing to long-term membership, or whether withholding these details would amount to denying the PPI Contributor the opportunity to make a fully informed decision about their participation. Negative phrasing, or priming, has been shown to produce more adverse responses to questioning^35^ which supports the concerns raised at the workshop.

A compromise was reached in which explicit information was written explaining who to speak to with concerns or difficulties in fulfilling the role of PPI Contributor, on the condition that this nominated PPI Lead would, if asked, provide all the required information on how to resign from the committee. This avoids negatively framing the commitment desired from PPI Contributors without actively withholding information. As only one piece of induction material provided through the unpublished resources discussed withdrawal from a committee, it was decided that this information is not considered a requisite topic to be covered in an Induction Pack but that CTUs should have information on the processes available to PPI Contributors if requested.

### Underpinning Theory

Academic theory has been used to support and justify the ordering and content of the eleven sections of the final document, in particular Anderson’s revision of Bloom’s taxonomy^26^ and Grow’s Staged Self-Directed Learning^36^.

It has been argued that the formal training of PPI Contributors negates the true lay perspective of a patient, carer or member of the public involved in clinical research. Ives et al.^37^ claim that untrained representatives of patient groups can make substantial, specialist contributions before and after clinical research is performed (for example when applying for funding or disseminating results). However, the skills and knowledge needed to support and oversee the conduct of research requires a level of training that would minimise the ‘outsider’ or ‘lay’ perspective. This paper, however, contradicts Ives’ argument and demonstrates that, as shown in Figure 3, the thorough induction and training of PPI Contributors is essential in providing the lower levels of topic understanding required to critically appraise study information and evaluate the data from a patient’s perspective. Comprehension of jargon and research processes is vital if a PPI Contributor is to provide insight and suggestions that would maintain medical and scientific integrity^38^.

**Figure 3:**
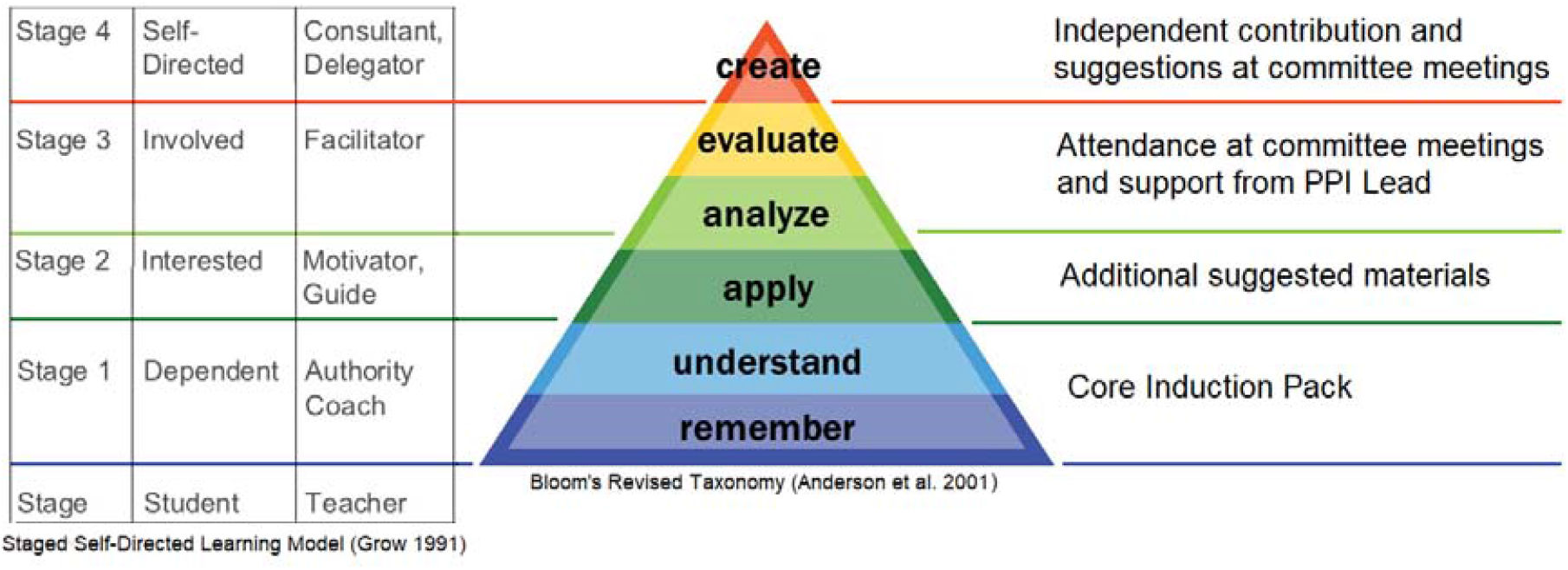
Application of Grow and Anderson theory to PPI Contributor training

In the production of this material, an assumption has been made that the reader of the Induction Pack is interested in understanding research concepts. Nevertheless, it was strongly stressed at the workshop that the entire document must be written in plain English as this could be the first exposure to technical language. This view is supported by the fact that almost 44% of the British adult population have a maximum reading comprehension of Level 1 or below which equates to that of a 12-13 year old^39^.

Accessibility of the Induction Pack has been enhanced through methods such as varying the layout of the text, using boxes to highlight key concepts and writing in short, active sentences in accordance with guidance issued by the Plain English Campaign^40^. Educational materials that have been created in collaboration with learners have been found to be more successful for self-guided learning^41^ therefore the re-review of the resource by the workshop participants provides assurance of the relevance of the document to its intended audience. An active effort has been made to avoid overwhelming new PPI Contributors with information, as this is a known deterrent for successful PPI^42^. The information layering and staged approach to learning allows the speed at which the topic is explored to be specific to each individual.

### Limitations

Although this research has gathered resources and information from a range of settings for primary and secondary research, including both published and grey literature, it has limitations.

Only three examples of induction materials were received from research charities, and this gave a limited view of what materials are currently in use. However, it is worth noting that most charities do not manage or coordinate clinical trials themselves: they do not have the clinical governance structure to oversee what could be complex interventions and therefore perform more patient-focused research rather than trials. The organisations approached may not have felt that they had relevant information to share. A wider representation of CTUs would also have added to the baseline knowledge that fed into this project but, considering that most of those who did respond sit on the UKCRC CTU PPI Sub-Group, it could be presumed that the documents provided came from units with the most interest in PPI and therefore the most developed resources.

Additionally, the success of the Induction Pack will be dependent upon trial teams completing the template to a high standard and distributing it to their PPI Contributors. Advice is provided to researchers to guide the completion of study-specific details; however, all additional information will still need to be written in plain English to ensure it is suitable for its audience.

### Relevance of the Induction Pack Template

The template created from this research has filled a gap in training resources for new PPI Contributors and has been made available to other CTUs to personalise and implement for their own studies and settings. The value of this role on trial oversight committees has thus far been underreported. It is hoped that through providing a template Induction Pack for CTUs more researchers will be encouraged to increase their PPI activity at committee level and track and evaluate the impact that comprehensively trained PPI Contributors can have on trial governance.

## Conclusion

To our knowledge, this is the first study to identify the requisite elements of an Induction Pack for PPI Contributors sitting on trial oversight committees. It provides a starting point for documenting the creation of evidence-based training materials which have been shown to be lacking in this field^25^.

The Induction Pack created as a result of this research is supported by current practice evidenced through published and unpublished literature alongside PPI review. The template demonstrates the wide range of topics to be considered essential when training lay members to join highly specialised committees which discuss very niche and jargonistic topics.

Further research should be conducted to create complementary guidance for researchers welcoming new PPI Contributors to trial oversight committees. This would support the development of more productive working relationships between PPI Contributors and researchers which has been identified as a top priority for future methodological research into PPI^33^. The role of PPI Contributors on oversight committees is under-represented in the literature and further research (such as the expansion of Daykin et al.’s study of relationships in oversight committees^43^) is eagerly anticipated.

The final Induction Pack created can be found online at https://www.ctu.mrc.ac.uk/patients-public/patient-public-involvement-resources/papers-guidance-templates/

## Data Availability

n/a

https://www.ctu.mrc.ac.uk/media/1416/induction_pack_for_ppi_contributors_on_trial_oversight_committees.pdf

## Endnotes

^a^ The TSC starter kit was unlocatable on the website and therefore could not be reviewed. It is, however, referenced several times throughout the website. It has been assumed that a link or landing page is missing.

## Contributorship

ECP conducted the literature review and research to create the draft Induction Pack under the supervision of BH and CDT, both of whom provided critical revisions. PB, JG, PH, RO and RS participated in the workshop as experienced PPI Contributors on oversight committees and provided review and edits to the final manuscript.

## Acknowledgements

We would like to acknowledge and thank the attendees of the Induction Pack workshop, the PPI Group at the MRC CTU at UCL, PPI leads from the UK CRC CTU Network and the Charities Research Involvement Group for their input and provision of resources which led to the creation of the Induction Pack. ECP’s affiliation changed from University College London to Imperial College London during the production of this manuscript.

---

Table 1: Search strategy employed for literature review

Table 2: Frequency with which topics appeared in unpublished induction/training materials provided by CTUs and research organisations

Table 3: Summary of feedback and discussions on draft induction pack from the workshop

Figure 1: Amended Talk-Estimate-Talk Delphi approach to PPI Contributor feedback on the induction pack

Figure 2: Chapter orders and summary of contents of revised induction pack

Figure 3: Application of Grow and Anderson theory to PPI Contributor training and how the induction pack supports active engagement in committees

